# Quantifying the contributions of asymptomatic and symptomatic colonized patients to *Clostridioides difficile* acquisition in oncological units

**DOI:** 10.64898/2026.05.08.26352751

**Authors:** Savannah Curtis, Mary M. Lee, Tiffany Hink, Kimberly A. Reske, Emily Struttmann, Zainab Hassan Iqbal, Candice Cass, Margaret A. Olsen, Sankalp Arya, Carey-Ann Burnham, Suzanne Lenhart, Erik R. Dubberke, Cristina Lanzas

## Abstract

**Objective:** Leukemic and hematopoietic cell transplant patients have one of the highest incidences of *C. difficile* infection (CDI). While CDI patients are considered the primary source of transmission, asymptomatic colonized patients (AC) can progress to CDI or contribute to in-unit transmission. We aim to quantify the roles of CDI and AC patients in *C. difficile* importation and transmission within oncological units.

**Design:** Prospective cohort study

**Setting:** Two leukemia and HCT transplant units in a large tertiary care hospital in the US

**Methods:** We developed a stochastic, individual-based network model to simulate *C. difficile* acquisition and transmission. Data from cultures and nucleic acid amplification testing (NAAT) obtained at admission and weekly, and toxin enzyme immunoassay (EIA) tests used for CDI diagnosis were used to calibrate the model. Healthcare worker room assignments informed the network structure. Key parameters were estimated via particle filtering.

**Results:** The model reproduced observed weekly test counts and transmission pairs. AC patients were the primary source of new colonizations: 51% were due to importation (of those, 88% were admitted as AC), and 49% were due to transmission (AC was the source in 92% of transmissions). Sensitivity analysis showed that these findings were most influenced by the colonization rate and rates of environmental contamination and cleaning.

**Conclusions:** These findings reinforce the role of AC, particularly via admission importation, in sustaining *C. difficile* transmission in high-risk hospital settings. Infection control focused on CDI effectively reduced onward transmission, as indicated by CDI’s low contribution to new colonizations.

## Introduction

*Clostridioides difficile* is one of the most common healthcare-associated infections in the United States. ^1^ The estimated national burden of healthcare-associated *C. difficile* infections (CDI) remained high, with 235,700 estimated cases in 2017. ^2^ Patients who undergo hematopoietic cell transplant (HCT) and chemotherapy for leukemia have one of the highest incidences of healthcare-associated CDI. ^3^ Such patients have near-ubiquitous antimicrobial exposures, immunosuppression, and a high frequency of prolonged hospitalizations.^3,4^ To improve CDI prevention in these patients, it is critical to understand their sources of *C. difficile* acquisition.

In HCT and leukemia patients, attribution of CDI acquisition sources is challenging. Their colonization with toxigenic *C. difficile* at admission is high, and during hospitalization, patients may experience intermittent carriage along with non-infectious diarrhea, complicating CDI diagnosis.^4^ Patients who test positive for toxigenic *C. difficile* on admission are up to 24 times more likely to develop a healthcare-onset CDI compared to non-colonizers.^5–7^ Patients can also become colonized and subsequently develop CDI during hospitalization. Both CDI and asymptomatic colonized (AC) patients with *C. difficile* are a source of pathogen transmission. CDI patients are often considered more infectious than AC patients, as they shed and contaminate their surrounding environments more extensively than AC patients. ^8,9^ However, AC patients outnumber CDI patients and are not targeted for transmission-blocking interventions. ^10^

The contribution of CDI and AC to new infections can be quantified using transmission models of *C. difficile* dynamics in healthcare settings.^11^ The majority of these models include AC and CDI patients as transmission sources, predicting that AC patients are likely a significant source of new colonizations. ^11^ However, there is considerable uncertainty in the predictions because models are often calibrated using only diagnostic tests performed on suspected CDI cases, as testing to identify AC is rarely performed in clinical practice. ^12^ Here, we aim to quantify the roles of CDI and AC patients in *C. difficile* importation and transmission within oncological units by integrating CDI diagnostic and active surveillance testing for AC patients into a transmission model of *C. difficile* acquisition. Specifically, we developed a modeling framework that embeds a transmission model of *C. difficile,* along with an observational model that simulates patient testing and sampling frequency, so that sparse testing and diagnostic tests’ characteristics can be accounted for.

## Methods

### Study Design and Data Sources

A prospective cohort study on *C. difficile* carriage was conducted in two leukemia and HCT transplant units at Barnes-Jewish Hospital in St. Louis, MO, between January 19 and July 22, 2019. Patient demographic characteristics are summarized in Table 1. Stool and rectal swabs for *C. difficile* culture and nucleic acid amplification testing (NAAT) were collected at enrollment and weekly until ward discharge. Of the 421 unique patients admitted to the study units, 384 (91%) consented to testing. Additional data were collected from the electronic records, including patient admission and discharge times, room assignments, medications, and laboratory tests. Clinical testing was triggered by symptomatic presentation and consisted of enzyme immunoassay (EIA) test for *C. difficile* toxin (Alere TOX A/B II). We also collected healthcare worker (HCW) staffing assignments. These data included shift-level records of HCW room assignments and were used to infer potential indirect contact networks between patient rooms. *C. difficile* isolates were also sequenced as part of a complementary study to evaluate the role of *C. difficile* strain diversity in colonization outcomes. ^13^ Environmental swabs were collected for *C. difficile* culture from bedrails, keyboards, and sink surfaces using three E-swabs (Copan). ^13,14^

**Table 1.** Patient demographic information for the 421 patients in the study units.

| Variable | Number of patients (%) |
| --- | --- |
| Enrollment ward |  |
| Acute leukemia (32 beds) | 353 (84%) |
| Allogeneic HCT (16 beds) | 68 (16%) |
| Length of stay (median and range) | 9 (0 - 152) |
| Age (median and range) | 63 (19 - 87) |
| Sex |  |
| Male | 249 (59%) |
| Female | 172 (41%) |
| Antiviral exposure | 348 (83%) |
| Antifungal exposure | 242 (57%) |
| Antibiotic exposure | 346 (82%) |
| History of CDI prior to enrollment | 28 (7%) |
| Graft vs. host disease | 15 (4%) |

To transform the testing data into a format suitable for comparison with model simulations, we assigned each patient’s infection status as susceptible, AC, or CDI on days when at least one test result was available to support the classification. Patients with a positive EIA test were considered CDI-positive. If a patient tested positive by NAAT or culture and had either an oral vancomycin start date or a CDI diagnosis date within 2 days following the positive test, they were also considered CDI-positive. Conversely, if a patient tested positive by NAAT or culture but lacked a positive EIA test or corresponding treatment or diagnosis date, they were classified as AC. Patients with only negative test results were classified as susceptible. If both positive and negative results were obtained on the same day, the patient was considered positive. Testing coverage was limited. Across all patient-days in the cohort, only 10.3% included at least one *C. difficile* test. Furthermore, because negative EIA results rule out symptomatic infection but provide no information about asymptomatic colonization, we could assign infection status for only 9.1% of patient-days.

### Transmission and Observation Models

We developed a stochastic, individual-based network model to simulate *C. difficile* acquisition and transmission. Each patient was tracked individually over time, and the model accounted for patient infection status, environmental contamination, and HCW-mediated interactions (Figure 1). Each patient was assigned an infection status that could change dynamically among three categories: susceptible (S), AC, or CDI. Environmental contamination was tracked in each room and represented all *C. difficile* exposure sources accessible during care, including contamination associated with patient contact (e.g., skin, bedding, frequently touched surfaces), rather than only persistent contaminated fomites. AC and CDI patients were assumed to shed *C. difficile* at equal rates and were modeled as continuously contaminating their environments upon colonization or infection. We performed a preliminary sensitivity analysis using an alternative 30:1 shedding ratio for CDI vs AC; this assumption did not substantially alter model behavior (see Supplemental Material). In the simulations, when a patient contaminated a room, their identity and infection status were recorded along with the time of contamination. If a patient contaminated the same room more than once, their most recent infection status was recorded.

**Figure 1.**
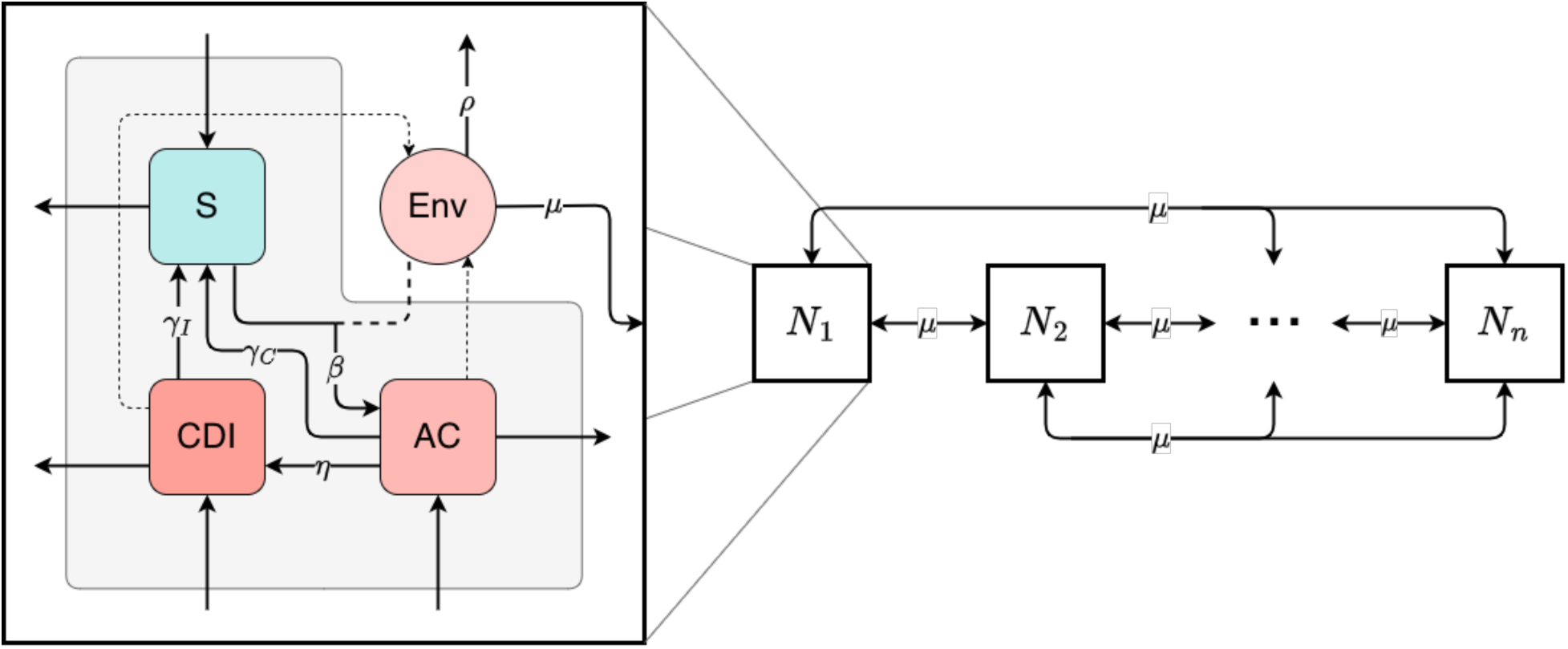
A visualization of the stochastic transmission network model. The right side of the figure depicts individual patient rooms (*Nᵢ*) and their connections through *μ*, which represents the rate at which healthcare worker movement transfers contamination between rooms. The left side of the figure shows the possible patient infection states (S for susceptible, AC for asymptomatically colonized, and CDI for *C. difficile* infection), along with the room’s environmental contamination level. Environmental contamination encompasses all sources of *C. difficile* exposure accessible during patient care, including transient patient-associated contamination, such as spores on skin, bedding, or frequently touched surfaces. Within each room, several events can occur: *β* for colonization, *η* for symptom development, *γ* for recovery or decolonization, and *ρ* for cleaning or decontamination. Events that involve interactions between the patient state and the room’s environmental contamination are indicated with dashed lines.

The model was embedded in a dynamic contact network informed by HCW staffing patterns. All patient movement and occupancy were modeled at the room level (i.e., admissions, discharges, and intra-unit transfers were defined with respect to specific rooms, not only the ward as a whole). Using room-level assignments for both patients and staff, we inferred connections between rooms based on shared HCWs. These connections formed a time-varying contact network, enabling contamination to spread between rooms through HCW movement. Patients transferring between rooms during their stay could further contribute to environmental spread. Contaminated rooms, in turn, posed a risk of transmission to newly admitted susceptible patients.

Each contamination event included a record of the source patient and their infection status, enabling indirect patient-to-patient transmission and linking transmission pairs. Transmission was defined as the occurrence of a new colonization event, which could occur when a susceptible patient occupied a contaminated room and became AC. When colonization occurred, the model assigned an attribution source by randomly selecting from the prior patients with contamination still present in that room at that time.

Transitions in patient status followed stochastic rules based on the duration of the incubation and infection periods. The model operates in continuous time, with transitions and events occurring probabilistically based on rate parameters. Patients could progress from S to AC, and from there develop symptoms consistent with CDI. AC and CDI patients could recover from either state. Routine daily cleaning was modeled as an imperfect decontamination process, with its effectiveness estimated during model calibration. Terminal cleaning following patient discharge was also included.

We implemented an observation model to translate predicted patient statuses into observed positive test results, accounting for testing frequency and test characteristics. For CDI patients, diagnostic testing using EIA was triggered by the onset of diarrhea and a suspicion of infection. The observation model incorporated the actual sample collection dates for each test and accounted for diagnostic test performance by considering both false positives and false negatives.

### Model Parameterization

We informed model parameters through a combination of literature review, institutional protocols, and data-driven inference. The following parameters were fixed based on existing empirical studies or clinical knowledge: the infectiousness/shedding ratio between CDI and AC patients, the success probability of terminal cleaning, and the probability that the observation model selects a CDI patient for EIA testing when one is present.

Because all movement was modeled at the room level, the timing of patient admissions, discharges, and room transfers, as well as test sample collection dates, were extracted directly from hospital records and used to define patient room-stays and testing schedules. The probabilities of being AC or CDI-positive upon admission were estimated by examining the results of all tests ordered within 48 hours of room admission.

Parameters related to EIA test sensitivity and specificity for CDI patients were drawn from published estimates.^15^ The model assumes that NAAT and culture are imperfect; therefore, their sensitivity and specificity are not 100%. For AC patients, the sensitivity and specificity of NAAT and culture were estimated using the 2-sample, 2-population Hui–Walter method as implemented in the *runjags* package in R^16^, with populations defined by unit and assuming independence between tests.

Other parameters were not directly measurable and required iterative estimation. We used an augmented particle filtering approach to estimate six key parameters that influenced the model’s fit to observed weekly test counts (Figure 2). ^17,18^ These included the rate of colonization from contaminated rooms (*β*), the rate of progression from asymptomatic to symptomatic infection (*η*), the duration of AC and CDI shedding (*γ*_C_ and *γ*_I_), and the rates of environmental HCW-mediated contamination (*μ*) and decontamination (*ρ*) through cleaning (Table 2). Full prior specification is provided in the Supplementary Materials.

**Figure 2.**
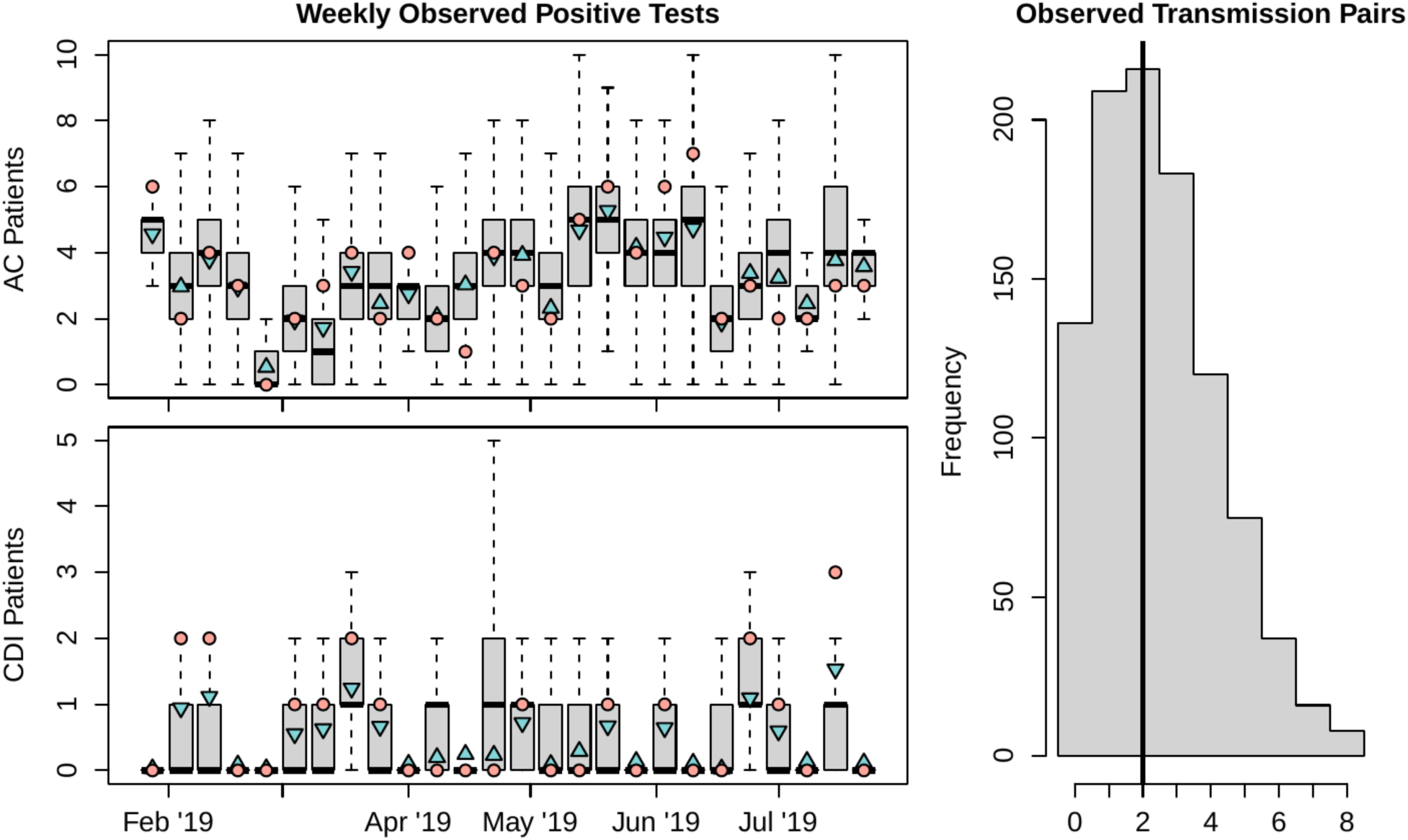
On the left, boxplots of weekly observed positive tests from AC and CDI patients for 1000 simulations from the calibrated model are shown. The pink circles in these figures are the data. The blue triangles represent the weekly weighted means from the observation model. Upward-pointing triangles indicate that the model overestimates the number of AC or CDI patients compared to the observed data, and model underestimation is indicated with downward-pointing triangles. On the right, a histogram of observed transmission pairs generated from 1000 simulations from the calibrated model is shown. The black vertical line corresponds to the two transmission pairs identified using whole-genome sequencing ^13^.

**Table 2.**
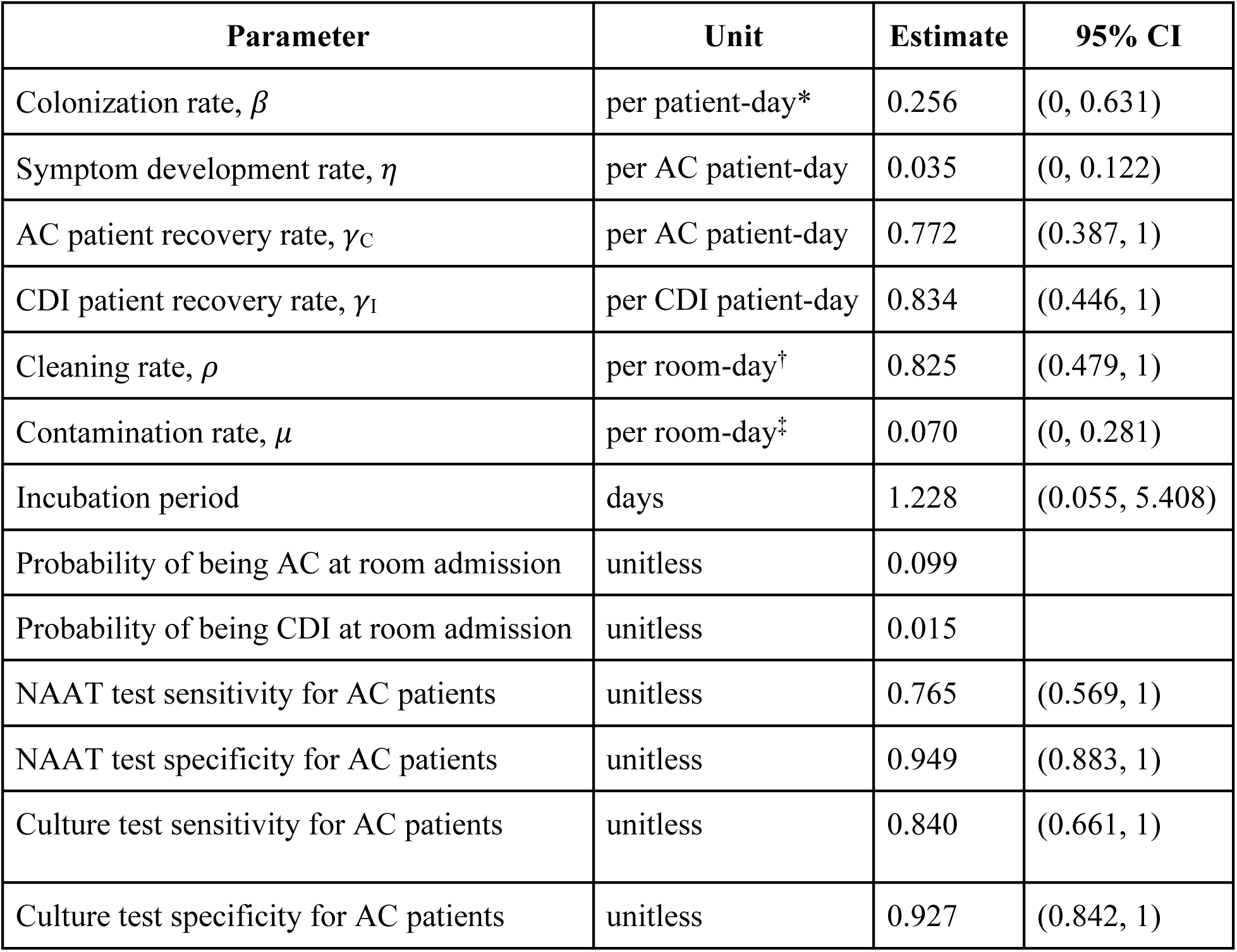
Parameter estimates for key model parameters. The six model rates obtained from particle filtering are shown first, followed by parameters estimated directly from data. Point estimates from particle filtering are taken to be the posterior mean, and the 95% CI is the 95% interval. Point estimates for test sensitivity and specificity are taken to be the medians, and the 95% intervals are the corresponding prediction intervals. *The colonization rate unit is per patient-day where the patient is susceptible and in a room exposed to *C. difficile*. †The unit for the cleaning rate is per room-day where the room is occupied and contaminated. ‡ The unit for the contamination rate is per room-day where the room is occupied, uncontaminated, and shares at least one healthcare worker with an occupied room that is contaminated.

We applied particle filtering to dynamically calibrate the model by simulating 1,000 parameter sets and comparing predicted weekly counts of positive tests with observed data. At each weekly time step, parameter sets that fit the data less well were discarded, whereas those that fit it more closely were retained and resampled. Once the filtering algorithm converged, we obtained posterior distributions for the six estimated model rates (Figure 3). Convergence was defined as successive parameter estimates remaining within the same order of magnitude, with no more than a one-digit change in the leading significant figure, or after three iterations had been completed. A complete list of parameter values, prior distributions, and estimated posteriors is provided in the Supplementary Materials. The code is publicly available at GitHub: https://github.com/lanzaslab/cdifficile

**Figure 3.**
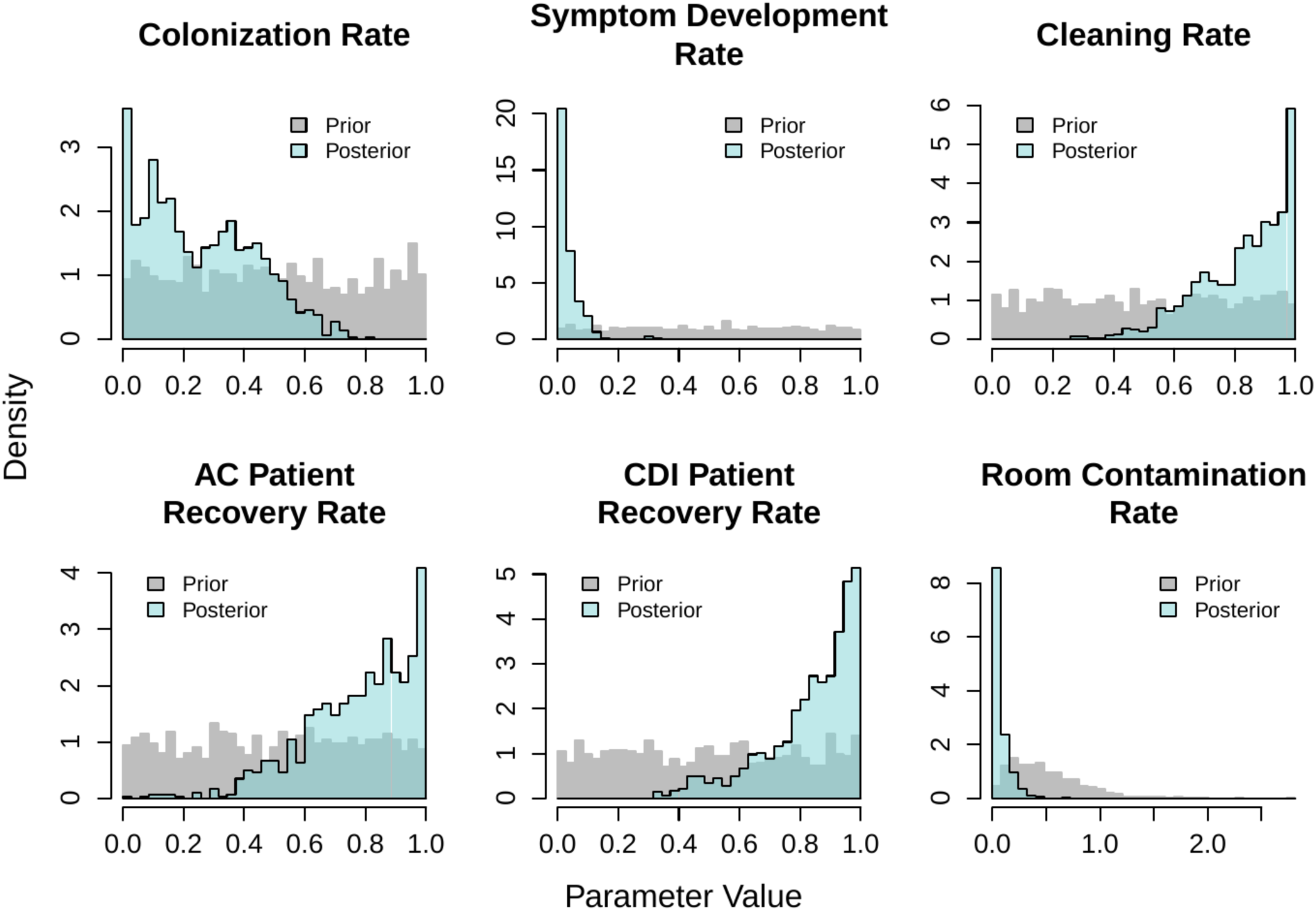
Histograms of the posterior distributions obtained for unknown model parameters after particle filtering. The prior distributions are shown in gray for comparison.

## Results

The model closely reproduced the weekly number of positive tests among AC and CDI patients (Figure 2). We also compared the number of simulated transmission pairs with those identified through whole-genome sequencing analysis.^13^ The model-inferred and empirically observed transmission pairs overlapped (Figure 2). The model parameters inferred from the study data, along with their mean values, are listed in Table 2. The posterior distributions for the parameters estimated using particle filtering are presented in Figure 3.

We simulated the calibrated model for 1,000 replications and averaged the source attribution for new AC and CDI cases over the 6-month study period (Figure 4). Figure 4A summarizes the attribution of all new AC and CDI; about half were due to importation into the room as AC or CDI. For acquisitions via transmission, AC was assigned as the source in approximately 90% of transmissions. When considering only new colonization events that progressed to CDI, the vast majority (87%) originated from importations (Figure 4B). Within importations, most were admitted already as CDI patients. The rest of the importations were AC patients who transitioned to CDI while in the unit. When unit transmission was responsible for CDI cases, the transmission source was an AC patient in more than 90% of the transmission events. The ranges for the source attribution are large because calibration relied on testing data from only 16 incident CDI cases over the 6-month study period. Therefore, to assess the robustness of these findings, we conducted a global sensitivity analysis focused on the average number of new colonizations per week across the four modeled acquisition pathways (see Supplementary Materials). The analysis indicated that new colonizations were most sensitive to the rate of colonization from contaminated rooms and the rates of environmental contamination and cleaning. These parameters influenced the persistence of contamination and the likelihood of indirect transmission. Other parameters had minimal influence on source attribution outcomes.

**Figure 4.**
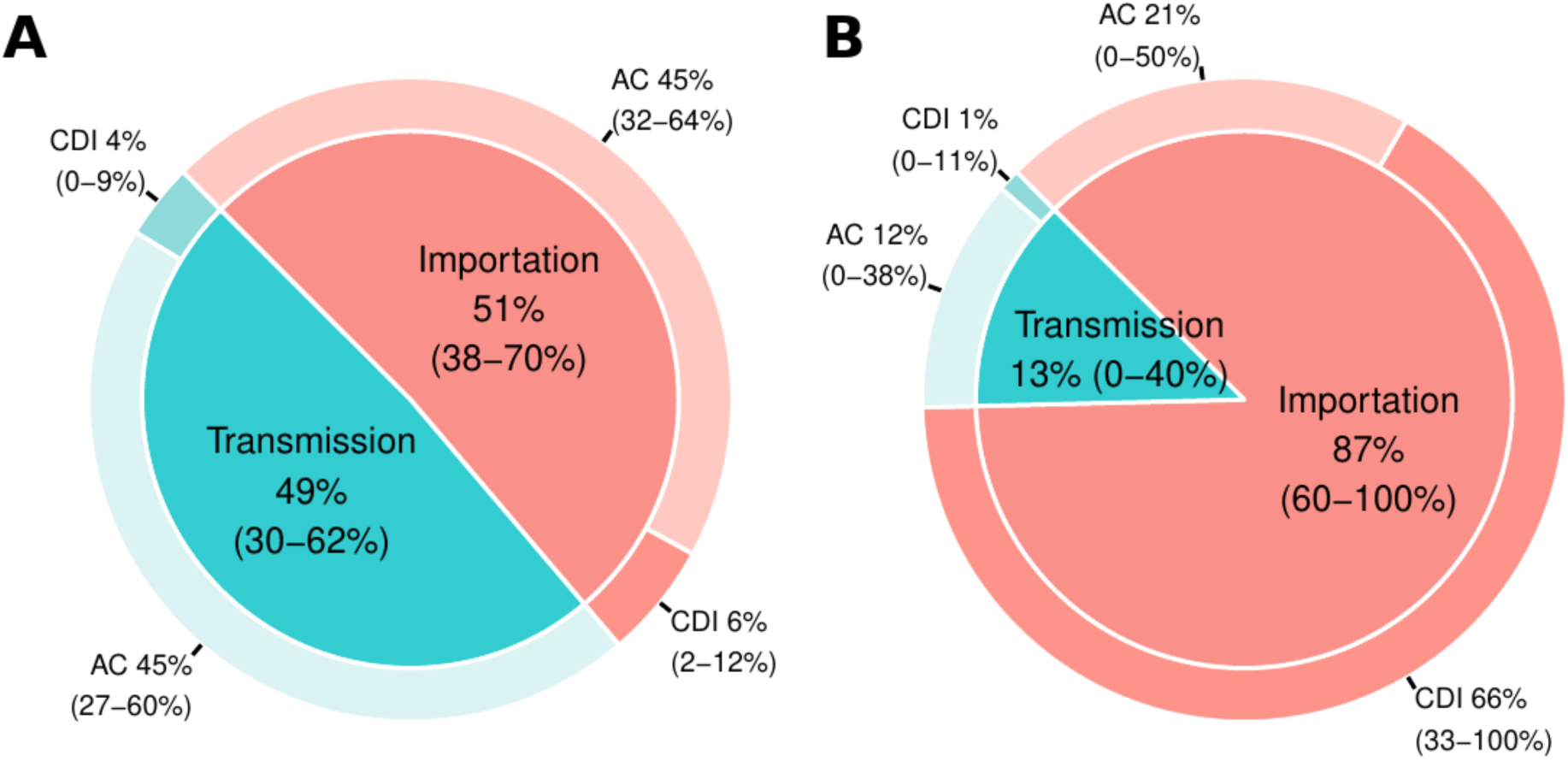
Pie chart for the source attribution of newly identified AC and CDI to ward transmission, with an AC or CDI as the source, or importation (i.e., the patient was already admitted to the room as AC or CDI). All model-predicted AC or CDI patients, both observed and unobserved, are included. Subfigure A shows the attribution proportion for all newly identified AC and CDI. Subfigure B shows only the subset of those admitted as CDI or progressing to CDI during the room stay.

## Discussion

This study differentiates the roles of AC and CDI patients in the acquisition and transmission of *C. difficile* in oncological units. We integrated testing data with a modeling framework that explicitly accounts for testing sparsity to quantify sources of *C. difficile* acquisition. Our model closely reproduced the number of positive tests and observed transmission pairs (Figure 2), indicating that the model captures the frequency and detectability of transmission events, even under low sampling intensity. Comparing the results from the transmission model with those of the observation model, which accounts for testing frequency and test characteristics, showed that a large percentage of predicted AC patients and transmission pairs were not detected through testing. The transmission model estimated 3.9 new colonization events per week, whereas the observation model indicates that only 0.9 (23%) of these were observed. This highlights an important challenge in quantifying the acquisition and transmission of healthcare-associated pathogens—acquisition events are only partially observed.

Considering only colonizations that progress to CDI yields a different picture of acquisition dynamics than including both AC and CDI patients. We found that room importation and transmission contributed similarly to new colonizations, and AC were the main contributors through both (Figure 4A). However, when only patients who progressed to CDI were considered, the majority of CDI cases were imported, either as CDI patients at room admission or AC at room admission and subsequent progression to CDI (Figure 4B). Similar to our results, Bruminhent et al. ^19^ traced the CDI cases in patients undergoing HTC to patients already admitted as AC, rather than to within-unit transmission. They used EIA to screen for AC at admission rather than NAAT testing, which may have led to an underestimation of AC at admission and subsequent transmission. ^19^ Overall, results indicate that although transmission from AC led to new AC colonizations (Figure 4A), CDI cases mainly developed in patients with *C. difficile* colonization already present at admission. These results, combined with the fact that our estimates for the recovery rates of AC and CDI patients indicated a short duration of *C. difficile* shedding (<2 days), suggest that patients transition quickly between multiple positive and negative test results, and that the majority of *C. difficile* new positive tests in the unit likely reflect transient carriage. ^20^ This carriage may have additional implications, as the AC patients could develop CDI after discharge. This work reinforced the need to consider strategies to prevent the transition from AC to CDI. Prior work in other types of units has also shown that within-unit transmission does not fully explain observed *C. difficile* cases ^7,21^, and thus interventions that prevent disease in AC can potentially reduce hospital-onset CDI cases ^22^.

We estimated key epidemiological parameters for *C. difficile* dynamics (Table 2 and Figure 3). Our estimate of the admission probabilities for AC patients aligns with prior estimates for this patient population. ^23,24^ Overall, patients with leukemia or undergoing HCT have a higher AC prevalence than other patient populations. ^25^ We estimated the sensitivity and specificity of the NAAT and culture tests for AC (Table 2). For AC, prior work has evaluated the diagnostic characteristics of NAAT tests using toxigenic culture as the reference method. ^26^ However, variability in enrichment broth, selection media, and incubation times can affect the sensitivity of culture tests. ^27^ The culture test had greater sensitivity than the NAAT test for detecting AC, while the NAAT test had greater specificity. Overall, both tests have relatively high sensitivity and specificity to detect AC.

The estimated colonization rate suggests an average of 4 days between a *C. difficile* exposure and acquisition. Once a patient is colonized, the transition rate from AC and CDI is 0.035 per AC-patient day, corresponding to a mean transition time of approximately 28 days. However, in the model simulations, most AC individuals did not transition to CDI. Considering only patients who transitioned to CDI reveals that the incubation period was typically short (<2 days) (Table 2).

Our findings align with early work on *C. difficile* acquisition, which indicated that the transition from AC to CDI was infrequent but, when it occurred, it did so shortly after colonization. ^19,28,29^ Curry et al. estimated that the median incubation period was 6 days for patients with prior negative cultures within 14 days, with an interquartile range of 3 to 12 days. ^30^ Our shorter incubation period estimate likely results from reduced gut microbiota diversity and pronounced gut dysbiosis, which are often present in these patients due to extensive antimicrobial exposure. ^30,31^ We also estimated the recovery, contamination, and cleaning rate parameters. The rate of routine room cleaning was estimated to be 0.825 per room-day. Assuming most hospital rooms are cleaned once per day, as reported in the protocol, this estimate can be interpreted as the probability that rooms are successfully decontaminated from *C. difficile* during routine cleaning. The 95% central interval for this estimate was wide, indicating that our data were unlikely to be informative about this parameter. The rate of room contamination via HCW movement was determined to be 0.07 contaminations per room-day. This can be interpreted as contaminated rooms being responsible for a subsequent contamination about every 14 days.

The main strength of the study is that we use both diagnosis and active surveillance data to fit the model and quantify the contributions of AC and CDI patients to *C. difficile* transmission and importation pathways. This study was conducted in a single healthcare facility; therefore, our parameter estimates and conclusions regarding the acquisition and transmission of *C. difficile* may not generalize to other settings or patient populations. Nonetheless, the modeling framework is readily adaptable to comparable datasets and can support future efforts to discern colonization, progression to symptomatic infection, and transmission pathways in other patient populations. In conclusion, our findings imply that infection control focused on CDI effectively reduces onward transmission from CDI, as indicated by the low contribution of CDI to new colonizations. But, interventions targeting AC can further reduce CDI by either reducing onward transmission from AC or by reducing the progression from AC to CDI.

## Data Availability

Aggregated and simulated data are available along with the R code for the simulation model at https://github.com/lanzaslab/cdifficile

## Acknowledgements

We would like to thank Avi Sawhney, who contributed to this work as an undergraduate researcher, for his assistance in analyzing healthcare worker staffing data and generating the time-varying adjacency matrices used in the transmission model.

The study protocol was approved by the Washington University Human Research Protection Office (IRB #201810103).

